# Hospital Preparedness and Response Framework during infection pandemic

**DOI:** 10.1101/2021.06.28.21259630

**Authors:** Bikash Bikram Thapa

## Abstract

Coronavirus disease 2019 (COVID-19) has put an unprecedented burden to world health, economy and social life with possible long-term consequences. The velocity and mass of this infection pandemic had already overwhelmed every robust health care system in the world. The evidence pertaining to this novel infection pandemic is evolving, so are the challenges in terms of adequate preparedness and response. In this review, we enumerate the strategic and operational domains and build a functional framework for the management of hospital mass infection incidents due to COVID-19 and similar future pandemics. This functional framework could assist health policy maker and health care worker to implement, innovate, and translate preparedness and response to save valuable life and resources.

## Introduction

The world has been hit hard with COVID-19 pandemic causing death in millions.[1] There has been unprecedented strain on health care system due to increase patient volume, increase acuity, special care requirements and resource reduction.[2] COVID-19 has exposed the vulnerability of the current national and global health system and call for novel and coordinated response.[3, 4] Current Pandemic preparedness plan is based on pre-COVID pandemic outbreak response model or the models of mass causality incident. Current COVID-pandemic is beyond pre-defined level (level 6 or level IV) of mass casualty incident. [3, 5]. International organization and several countries have provided documents for hospital preparedness and emergency response frameworks for COVID-19 pandemic, but are not uniform.[6-8] Awareness and preparedness to prevent, detect and respond the pandemic are inconsistent among world countries.[9, 10] According to international health regulaiton annual report data, out of 182 coutries, only 50% have operational readiness capcities in place.[11] The key principles of the pandemic emergency preparedness and response are protection of vulnerable, functionality of the health care system, support to the staff, and treatment of the infected through multi-disciplinary multi-level approach.[12, 13] A cross-sectional survey in 102 emergency departments of 18 European countries documented a lack in early availability of adequate hospital preparedness and response plan.[13, 14] It is necessary that nations and hospitals innovate or translate the policies considering the local resources and capacity.[15, 16] Health care institute require strategical preparedness and response plan as a part of broader ‘infectious disease epidemic plan’ (IDEP) considering the cases and capacities.[17, 18]This review is aim to outline the framework that form the basis for hospital preparedness and response during hospital mass infection incident caused by COVID-19 pandemic.

## Materials and Method

The web-based (Medline and Google scholar) literature search was done with keywords; hospital preparedness and response in the title, and COVID-19 or SARS-Cov-2 in all fields. The research article retrieved through online search engine (August 2020) were reviewed by the author (October 2020 to April 2021) based on PRISMA protocol (Figure 1). The objective of this review was to identify the 1. Strategic domains 2. Operational activities and 3. Conceptual framework necessary for hospital preparedness and response to mass infection incident in the context of COVID-19 pandemic. The strategic domains were structural elements necessary to conceptualize hospital preparedness and response framework for patient care. Operational domains translates concept and design into action to achieve goals [19]. The hospital preparedness and response framework was designed based on the conceptual theory of “Resilience Framework for Public Health Emergency Preparedness” conceptualized by Khan [20] et al. In this conceptual framework all domains (strategic and functional) were fit together to coordinate effective preparedness and response plan leading into a resilient health system.

**Figure1.**
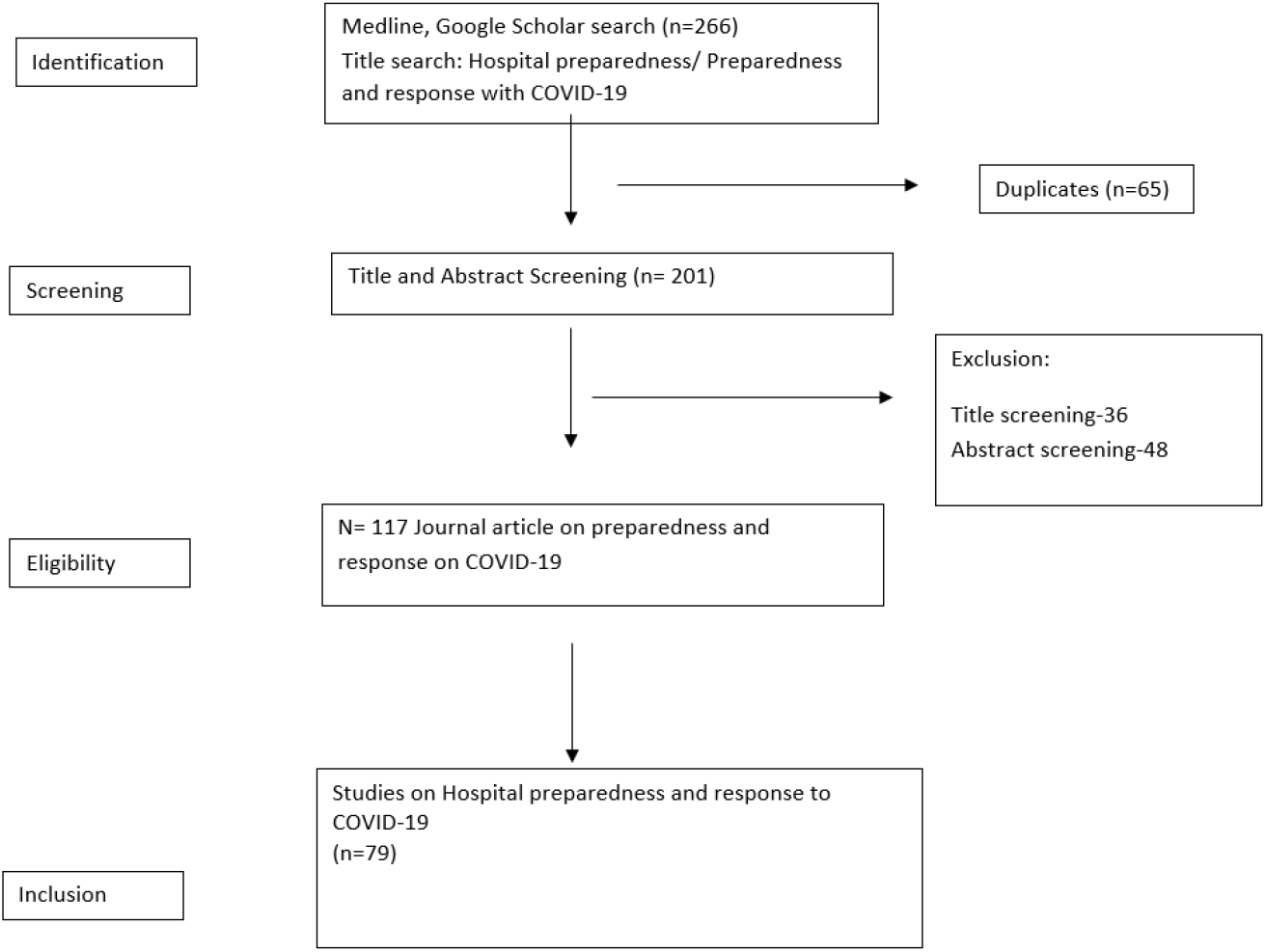
PRISMA Protocol for Review.

## Result

The hospital preparedness and response framework with built-in strategic and operational domains of hospital preparedness for infection pandemic is depicted by figure 2. The strategic domain for hospital preparedness for COVID-19 pandemic related mass infection incident were; 1. Leadership 2. Ethics 3. Clinical care 4. Resources 5. Infection prevention and control 6. Data management, which were bonded together in action by effective communication, training and education, and research (Figure 2-hospital preparedness and response framework for infection pandemic). An array of operational activities underscored the successful function of the structural framework. Leadership and governance were the platform for all strategies and operations. The elements of framework were integrated and protected with relevant ethical values and principles. Leadership was emphasized for accountable, transparent, efficient and effective management of health care system. Resources allocation was most challenging and controversial. Managing strained resources (staff, space and stuff) during pandemic requires surge capability. Quality clinical care, resource optimization, efficient data management, and infection prevention and control strategies, when translated into crisis standard ethical action help in realizing the goal of hospital preparedness and response to mass infection incident.

**Figure 2:**
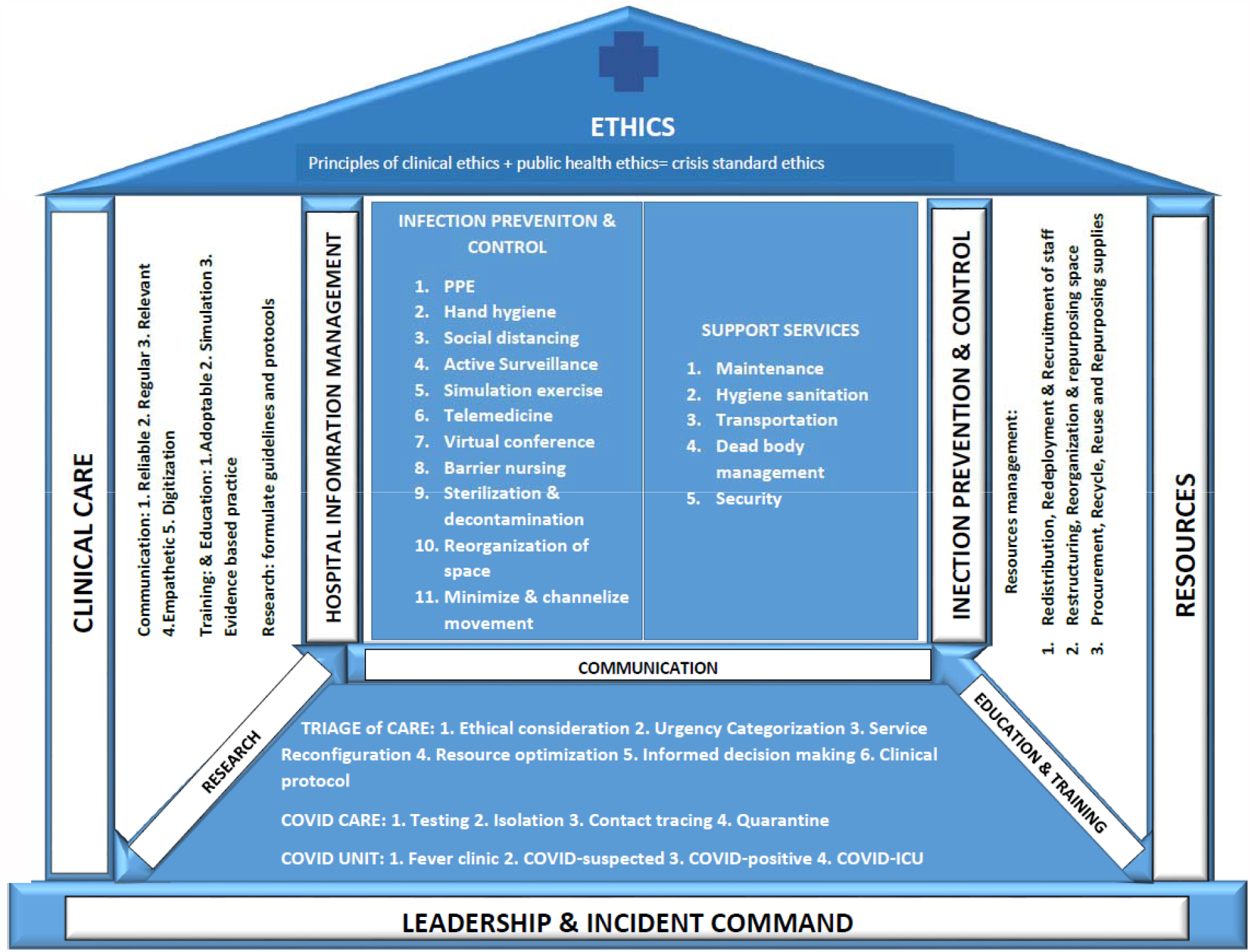
Hospital preparedness and response framework for infection pandemic.

## Discussion

Preparedness involves decision making to develop strategies to mitigate the health threats from disease outbreaks. The key activities of the policy planning are anticipation, assessment, prevention, preparation, response and recovery. Evaluation of the context, analysis of the risk, and acknowledgement of the stakeholders and resources is essential to plan hospital preparedness and response measures.[21] Non-uniformity of the context, difference in level of risk and variable stakeholder make hospital preparedness and response framework dynamic and complex. The strategic risk analysis of infection pandemic depends on disease epidemiology and surveillance (incidence, prevalence, and transmissibility), vulnerability (age, comorbidity, socioeconomic determinant of health) surge capacity, and coordination. This provides strategic tool for preparedness and response. [22]

Strategies and operational tools are instrumental to achieve preparedness and response. The resilience of the hospital preparedness framework constructed is based on the interconnections of the elements. The essential elements of the public health emergency preparedness framework conceptualized by Khan et al are; governance and leadership, ethics and values, resources, workforce capacity, communication, surveillance and monitoring, practice and experiences, learning and evaluation, and collaborative networks. The interlinking and the cross-cutting of the elements guaranteed the robustness of the framework. [20]

### 1. Leadership

Leadership and planning operationalize and optimize response effort.[2] Formation of central Incident Command Center with multidisciplinary multi-level team leaders is the first step towards hospital preparedness and response to COVID-19.[4, 23] Regular conference and discussion guide real-time solution. Transformational and occasionally authoritative leadership model are successful approach during health emergency. The leadership facilitate as a mentor for policy initiatives and implementation, through necessary strategies and operational guidelines.[23, 24] Farcas provide a model where hospital incident command is responsible for all sort of hospital activities during pandemic.[4, 7]. A effective command or leadership process through: situation reporting and assessment; problem identification and prioritization; solutions and new assignments; and monitoring and evaluation.[25] The pandemic management requires commitment for shared decision making and effective communication at each level and process.[26]

Physician leaders are at unique position to understand clinical challenge and can legitimately address the issues at strategic, peers and patient level with expertise and skill [27] The five cultural elements of medical leadership are; (1) clear vision, (2) vision aligned objectives, (3) Resource engagement and support, (4) Research, innovation and training, and (5) team work [28, 29] The success of leadership depends upon understanding the situational factors. The task oriented authoritarian style is recommended in emergency or critical environment. Medical emergencies multidisciplinary multilevel job where the chance of un-coordination and contradiction remains high. Overlapping and duplication of strained resources can happen. Leadership moderates willingness to take responsibility and act with maturity and emotional intelligence. Leaders obtain authority from knowledge of situation (internal and external), disease, and the process.[30]

### 2. Ethical Consideration

Ethical consideration provide leadership with principles and tools to achieve goals. Resource allocation, care-triage, service reconfiguration, duty to care, information dissemination, and research all need ethical consideration to run effective and efficient health care system during disease pandemic. [31]The four fundamental principles of ethical health care are autonomy, non-maleficence, beneficence and justice. However, scarcity of resources and cancellation of non-urgent cases conflict with principle of clinical care ethics. Trading infection prevention and control measures, and forced restrictive measures with rising psychological impact and economic consequences necessitates the shift of clinical ethics to public health ethics.[32] Family and public health oriented informed decision making preserve both the individual and public rights for health care during crisis.[33] COVID-19 pandemic should bring social justice, equity, utilitarianism, reciprocity, and solidarity to forefront of health care management.[34-36] During crisis the equality is trade off with benefit. Resources are ethically diverted to save more lives or life years. “Sickest first and youngest first approach” rationalize crisis standard ethics.[37] Ensuring psychological wellbeing, occupational safety, and family care of health care providers ensure value proportionality of their service. The first come first serve, clinical protocol based admission, equity and reciprocity based selection, and clinical score (sequential organ failure assessment-SOFA) all facilitate triage decision during pandemic crisis.[38]

Both clinical and public health care grounded on fundamental ethical values need to operate with clear mechanism of transparency, accountability, effectivity and efficiency.[39] Narration of principle into crisis standard substantive and procedural ethics can legitimate health care activities during COVID-19 pandemic.[40]

### 3. Resources

Resources (supplies, space and staff) preparedness is necessary to meet the infection surge in hospital. Surge capacity with efficient and sustainable responses are the key denominators of resilient health care system.[41] Surge capacity is designed to re-activate the health system within 12 hours and sustain it for 2 weeks.[7] Adequate supplies of scared and expensive medicines, supplies, and life-sustaining equipment requires public-private collaboration, and solidarity. Reorganization and restructuring of infrastructure; redeployment and recruitment of personnel; redistribution and repurposing of the supplies are key operational activities to meet the increase demand of resource.[7, 42, 43] Resources supplies can be managed with “step-wise escalation” into conventional capacity, contingency capacity, and crisis capacity to maintain supply chain, and work teams.[42, 44] Resource strain and resource burning is rate limiting step of COVID-19 care outcome. A “Brick system” adopted and modified by Indian army medical corps has been a crucial model to manage inventory in health emergency, where bricks of different size is assigned based on different level of COVID care.[45]

Germany adopted COVID-19 rapid response infrastructure (CRRI) model to manage rapid escalation to maintain critical care services functionality.[46] COVID-care unit should have dedicated diagnostic and other support units in close proximity; one-way movement of resources and personnel; and negative pressure environment in all involved space.[13] Restructuring should ensure separate donning and doffing area for each COVID-care unit; dedicated resting and refreshment area; satellite pharmacy and supplies.[47] Deferring ‘Blind rehabilitation’ of vulnerable population and admission of the stable COVID patient can surge space. A tertiary care hospital in Italy produce 33% surge in health care provider and reduction of surgical cases to less than 10% through reorganization and service reconfiguration.[13, 23] The hospital support units for maintenance, hygiene and sanitation, transportation, security, and mass fatality management are essential life lines for hospital preparedness and response to COVID crisis. [4, 48]

Redistribution of resources is dynamic process.[49, 50] Internal redeployment and external recruitment can expand human resources. Knowledge and experience based group division and redistribution of health care worker for different working zone adds to resilience and engagement. Kichhar et al. adopted three division with resident doctors, nursing and supporting staff to serve 8 hour shift and cycle of 9 day rotation in ICU and 15 days rotation in COVID-isolation ward[51] In order to prevent HCW burnout it is advisable to ensure cooling-off time and or “buffer ward” (non-COVID) rotation in between.[47] Minimal risk exposure of vulnerable with maximal use of professional skills is necessary for efficiency and effectiveness of the health care system.[52, 53] Medical students are more suitably posted in screening area or fever clinic. COVID-19 duty shift and staffing ration need to justify the wellbeing of health care provider, the surge capacity, and equity in care.

### 4. Clinical care

The goal of health care center is to mitigate, plan, response and recover, the unexpected burden posed by COVID-19 pandemic. Care of COVID patient is multidisciplinary approach. The clinical care strategic includes treatment of COVID-19 case (from triage to critical care unit), infection control, prevention of nosocomial transmission, and elective/non-COVID case management. [7, 54] The biggest challenge to case management is novelty of the disease.[42] The four pillars of effective patient care are a) testing of suspects, b) Isolation, treatment and referral of confirmed depending upon severity, c) Contact tracing and screening, and d) quarantining of the close contacts.[18, 24, 53]. During pandemic the essential and emergency services need continuity to protect public and personal health.[18, 55, 56] The continuity care of non COVID patient need categorization (low, high, emergency and non-essentials) and service reconfiguration, based on urgency and impact of COVID-19.[41, 56-58]

Algorithm and guidelines rationalize the COVID case management. Anticipation, and preparedness for clinical care ahead of incident had favorable outcomes in hospital and critical care unit.[43, 59-61] Infectious disease society of America (IDSA), The European Centre for Disease Prevention and Control, and World Health Organization all provide strategies, criteria and technical guideline for testing, isolation, and treatment.[62-65] Sequential organ failure assessment (SOFA) triage protocol was in practice for prioritizing access to critical care during pandemic. SOFA score < 7 is given highest priority and 8-11 with intermediate priority. Score >11 can be either managed medically or palliative care or discharged.[38] Pandemic could cause surge in demand of palliative care preparedness and plan that might necessitates separate surge capability and contingency.[66] Occupational health protection protocol and use of PPE is done according to hazard risk level. Donning and doffing during COVID duty starts with hand hygiene and includes minimum of N95 face mask,/respirator two layers of surgical gloves, head cover, eye/face shields, full sleeve gown, apron and shoe cover[23, 53, 59, 67].

COVID and non-COVID cohort of patient should be managed separately to prevent and control nosocomial transmission of the disease.[47, 58, 68, 69] The diagnosis and triaged is based on epidemiological, clinical, and investigation findings. However, clinical features of the COVID-19 are non-specific and warrant strong association with travel and contact history. The sequence of pre-triage, triage, diagnostics, staging and definitive care are important to streamline the process and resources.[13, 69] COVID-care center requires Fever clinic, COVID-suspected ward, COVID-isolation ward, and COVID-critical care unit with robust laboratory and imaging facilities. RT-PCR test is highly specific test but the accuracy is yet to be evaluated in systematic analysis (false negative result as high as 40%). [70, 71] Pharmacotherapy and other therapeutics need careful selection based on evolving evidences and cost effectiveness. Most of the trials related to Hydroxychloroquine including “SOLIDARITY” trial was terminated owing to the lack of clinical benefit.[72] In comparison to placebo Remdesivir reduced time to clinical improvement (Hazard ratio for improvement is 1.23).[73] Dexamethasone had shown reduction in 28 day mortality in hospitalized patient when compared to usual care.[74] Attending health care provider requires “point care skill” (ultrasound, echocardiography etc) in ward and ICU.[47] Respiratory and hemodynamic support is crucial. Out of 72314 COVID-19 confirmed cases, Wu reported mild, severe and critical disease in 81, % 14% and 5% respectively with overall mortality of 2.3%. 19% of these patients had hypoxic respiratory failure who need invasive or noninvasive ventilation.[65, 75] Critical and informed decision making is necessary to justify limited scientific evidences and scarce resource (personal protective equipment, respiratory support and infection control means) utilization in mass critical care scenario.[57, 76] It is argued that a single consent mentioning all possible procedure at the time of admission avoid unnecessary conflicts and contacts.[47]

### 5. Infection prevention and Control

“Swiss cheese model” that provide multiple layers of defenses in prevention and control of infection is a safe hospital strategies.[77] A triple check system; help desk, screening desk and emergency evaluation desk are deemed necessary.[51] Hand hygiene, social distancing, and face mask were underscored as universal precautions for infection prevention and control (IPC). Personal protective equipment are recommended according to the risk of exposure in different level of work station (biosafety level). [53, 57] 48% IPC professionals reported shortage of PPE in a multinational survey study. It was as high as 64% in low income countries.[78] Strict precaution is recommended in area with high chance of aerosol producing procedures that includes sample collection, nebulization and intubation. Simulation based training effective means to make HCW conversant to new environment as well as for IPC. [79-81] Multimedia display regarding precautions and standard operating procedures reinforce preparedness and response.[47] Time spent with COVID-19 suspected and confirmed cases can be minimized using video-surveillance, in-room phones, and virtual visitors.[7] HCW visit to COVID-19 care unit should have time and frequency count per day depending upon the available human resources. Disposable unit of personal supplies and consumables are encouraged. All administrative, educational and clinical meetings when limited to virtual conference maintain social distancing for IPC. Emergency department, hospital wards, Operation Theater and Intervention center need rapid transformation in line with segregated and barrier treatment/surgery for any suspected or confirmed cases of COVID-19.[23, 47] Movement of resources, and COIVD and non-COIVD patient should not cross paths. Clear visual and spatial separation of care areas maintain barrier to exposure and spread.[23] Precaution in operation theatre includes proper decontamination, disinfection and sterilization of equipment and instruments[82]; undertaking surgeries based on newly adopted CVOID-pandemic guidelines; and judicious application of surgical techniques and equipment.[56] Institutional policy, installation of IPC facilities, knowledge, active screening and surveillance, and response all are key elements of infection prevention and control.[18, 24]. The reported rate of infection transmission to HCW ranges from 0-0.001% in India [47, 51], and 7-18% in Europe [83]. Mandatory screening of all visitors, visitor log, and maintain health care worker’s contact diary helps in disease surveillance. It is duty of system to equip and reciprocate health care workers for better response to pandemic. Self-reporting, surveillance, chemoprophylaxis, sanitation and isolation are measures of occupation health protection. [47]

### 5. Information Technology (Infotech)

The information management system and technology has multiple purpose including: secure and quality data management; effective communication; increase accessibility to health care; improved disease diagnosis and treatment. [84, 85] Electronic tools and data base provide evidence based preparedness and response to COVID-19 pandemic by early alert, case detection, contact tracing and follow up. World Health Organization has been a strategic partner to provide platform for digital tools.[86]The InfoTech and “infodemics” in COVID era is maintaining health and health care delivery. Digital media has been widely approved at national and regional level to provide updated resources, guidelines, education and awareness. It creates smooth functioning of the interconnected elements of the hospital preparedness and response. E-learning, telemedicine, tele-conferencing are viable alternatives for response activities. Pandemic related spatial and temporal data are widely available through interactive policy informatics platform and research repositories.[87]

### 6. Information and Education

Information is organized data or knowledge that provides a basis for decision making. Information is best resources available at hospitals.[88]Hospital information management system is a link between logistics and operations for effective health outcome. The channels should be reliable, regular, and relevant.[41] A focal person in incident command chain serve the purpose. Information sharing and clinical decision making need evidence based, empathetic, respectful and humanitarian approach.[89] Digital technology is not only the vital source of evidence based medical practice but also a smart means of active disease surveillance.[85, 87] Information sharing pertaining to reorganization of hospital services, infection control strategies, visitor’s movement restriction, family-informed decision making, clinical guidelines and protocols should be clearly communicated to all responsible stakeholders (in patient care).[41, 90] Education and training promote adaptability for emergency response and prepare for unknown future. Cycle of learning and evaluation foster improvements and innovation for better preparedness that are locally applicable and affordable.[20] Continue medical education and regular training is necessary to enhance evidence based health care during pandemic. Simulation based exercise has been proven effective for preparedness and response to novel pandemic. The “plan-do-study-act cycle” is a valuable process of simulation to adopt new challenges.[81, 91]

### 7. Communication

Communication is a strategy to network components of hospital preparedness framework by delivering clear and consistent message. The aim of robust communication is to influence health related behavioral change, promote cohesiveness in networking, and to promote professionalism in health care. Communication thrive in relationship, feedback, and collaboration.[20] Honest and effective communication of crisis and risk of pandemic builds trust, and credibility. Atuguba argue that effective communication is the essential social determinants of the mental and physical health in COVID-19 pandemic. [92] The key messages should be communicated timely, and reviewed regularly. Effective team work and communication is responsible for lowering hospitalization and costs; improve efficiencies; enhance service user satisfaction; increase staff motivation, productivity and innovation; reduce medical error and overall morbidity and mortality[93] Conversations require empathetic listening and responsive action to address the loss and grief. Conveying lack of resources, critical medical condition, death of patient require humanistic approach.[94]

### 8. Research

The medical practice and protocols are rapidly being reconfigured in order to generate knowledge to provide best of medical care. Research provide ethical approach for duty to care, duty to safeguard, and duty to guide the health care. Conducting medical research itself demands highest ethical and safety standards. Institutional and national health research governing body have crucial role to play.[95]Ethics for clinical trials is under scrutiny. There is growing argument to preserve rights of a research subject during randomized trials.[96] Research and trials must be informed by equitable access to innovation. Social and behavioral science researches are valuable to explore the impact of pandemic. Scientific research is the basis for present and future infection pandemic policy, preparedness and response. [97]

## Conclusion

The COVID-19 and its consequences are still evolving. This review provides comprehensive frameworks and principles towards hospital preparedness in wake of COVID-19 pandemic. Medical leadership, ethics in health care, resource optimization, quality clinical care, infection control and, information technology are the building blocks of hospital preparedness framework. The emerging scientific evidence will help to map the policy and strategy accordingly. The past experience with pandemic and disaster response need to be translated into more effective and efficient universal model to manage hospital mass infection incident. A single model might not be adoptable for all. The strategy to address the inequitable health care system across the world need further research and evaluation. COVID-19 pandemic and the mass infection incident is beyond any measureable scale. The global pandemic calls for global preparedness and response. The model presented in this review will supply policy maker a basis for adoptable and sustainable hospital preparedness plan to save lives, life years and optimize resources. Emergencies and crisis are unpredictable; better be prepared than perish.

## Data Availability

Data referred to the manuscript are available with the corresponding author

## Acknowledgement

none

